# Identification and Functional Characterization of Bicaudal-D2 As A Candidate Disease Gene in Autosomal Recessive Consanguineous Family with Dilated Cardiomyopathy

**DOI:** 10.1101/2022.02.03.22269864

**Authors:** Kai Luo, Chenqing Zheng, Rong Luo, Xin Cao, Huajun Sun, Huihui Ma, Jichang Huang, Xu Yang, Xiushan Wu, Xiaoping Li

**Affiliations:** Department of Cardiology, Sichuan Provincial People’s Hospital, University of Electronic Science and Technology of China, Chengdu, Sichuan 610072, China; Chinese Academy of Sciences Sichuan Translational Medicine Research Hospital, Chengdu 610072, China; Shenzhen Aone Medical Laboratory Co., Ltd., Shenzhen, China; Institute of Geriatric Cardiovascular Disease, Chengdu Medical College, Chengdu, People’s Republic of China; School of Acupuncture-Moxibustion and Tuina, Chengdu University of Traditional Chinese Medicine, Chengdu, China; Department of Pathology, Sichuan Provincial People’s Hospital, University of Electronic Science and Technology of China, Chengdu, Sichuan 610072, China; The Center for Heart Development, Hunan Normal University, Changsha, China

**Keywords:** dilated cardiomyopathy, *BICD2*, zebrafish model, RNA-seq

## Abstract

Familial Dilated cardiomyopathy (DCM) is a genetic cardiomyopathy with reduced left ventricle function or systolic function. Fifty-one DCM genes have been reported, most of which are inherited in an autosomal dominant manner, while those caused by a recessive manner are rarely observed. Here we identified an autosome recessive and evolutionarily conserved missense variant, c.2429G>A: p.Arg810His, in bicaudal D homolog 2 (*BICD2*), which was segregated with the disease phenotype in a consanguineous family with DCM. Furthermore, we confirmed the presence of BICD2 variants in 3 of 210 sporadic cases, highlighting its candidate causative role. Interestingly, we discovered that *BICD2* expressed higher in cardiomyocytes from DCM than that from control by reanalyzing published scRNA-seq dataset, implicating its possible involvement in cardiac function. We next explored *BICD2* function in zebrafish model at both embryonic and adult stages. Knockout of *bicd2* resulted in partial embryonic lethality in homozygotes, suggesting a vital role for *bicd2* in embryogenesis. We performed zebrafish echocardiography to detect indices of ventricular size at the adult zebrafish stage. Intriguingly, dilated hearts, decreased ejection fraction, cardiac output and stroke volume were observed, suggesting a phenotype similar to human DCM in *bicd2*-knockout homozygotes. Furthermore, RNA-seq confirmed the largest transcriptome shift in *bicd2* homozygotes. Gene Set Enrichment Analysis(GSEA) of *bicd2*-deficient fish showed altered gene expression enriched in cardiac pathways and mitochondrial energy metabolism. In conclusion, we found for the first time that an autosomal recessive *BICD2* variant is associated with familial DCM, providing further insight into the molecular pathological mechanism of DCM.

## Introduction

Cardiomyopathy is a heterozygous disease of the heart muscle, leading to heart failure. Cardiomyopathy mainly include dilated cardiomyopathy(DCM), hypertrophic cardiomyopathy(HCM), as well as arrhythmogenic right ventricular cardiomyopathy (ARVC)[1]. DCM is a cardiomyopathy characterized by dilatation of the left ventricle or bilateral ventricles with myocardial systolic dysfunction[2]. The prevalence of dilated cardiomyopathy reported in global is 1/250[3]. It is the third leading cause of heart failure and the most common cause of heart transplantation [4], but its etiology and pathogenesis remains elusive.

Familial DCM is now more commonly diagnosed, owing to the improved awareness of disease-causing genetic variants and genetic screening. Familial DCM is found in 20-35% of patients with DCM, and 80% of DCM patients inherited in autosomal dominant pattern, 10%-15% of DCM patients belong to autosomal repressive or X-linked inheritance[5]. Previous DCM studies unraveled >250 associated genes spanning >10 gene ontologies, suggesting a complex and diverse genetic architecture[6]. A DCM Gene Curation Expert Panel curated a final set of 51 genes proposed to have a monogenic role in isolated, idiopathic DCM in humans[6]. And 12 out of 51 genes, ie. *BAG3, DES, FLNC, LMNA, MYH7, PLN, RBM20, SCN5A, TNNC1, TNNT2, TTN, DSP* were classified as having definitive or strong evidence[6]. Other 7 genes including *ACTC1, ACTN2, JPH2, NEXN, TNNI3, TPM1*, and *VCL* were classified as moderate evidence[6]. Although assessing as having high evidence, these 19 genes explain only a minority of cases, leaving the remainder of DCM genetic architecture incompletely addressed[6].

Most reported DCM variants inherited in an autosomal dominant pattern[5-7], while autosomal recessive(AR) inheritance in familial DCM was less frequently observed and implicated in pediatric DCM[8-10]. Consanguineous families is ideal study objects for investigating AR DCM. However, few researches focused on familial DCM in consanguineous families[10-12].

Bicaudal-D2 (*BICD2*) is a dynein activating adaptor protein that plays a critical role in microtubule-based minus-end-directed transport[13, 14]. Mutations in the human *BICD2* have been linked to a spectrum of neuronal disorders, in particular to a dominant mild early onset form of spinal muscular atrophy[15-18]. The *BICD2* protein harbors a C-terminus recognizing and binding to the material being transported and an N-terminus containing three helix-helix structural domains that bind to the kinesin tail and kinesin-activating protein filaments to form a sandwich structure of the kinesin complex, which activates the kinesin and allows the kinesin complex to travel along the microtubule cytoskeleton to transport cargo[19]. *Bicd2*-KO mice showed significantly lower *LMNA* expression at protein level, decreased *MKL-1*/*SRF* activity, and significantly downregulated expression of its downstream α-actin, α-coactin and membrane association proteins, and microtubulin[20]. *MKL-1*, a member of the actin family, plays an important role in cardiovascular system development and function as a cofactor of serum response factor (*SRF*) that activates *SRF*-dependent transcriptional regulatory elements. In nuclear fibrillar protein A/C gene (*LMNA*) knockout mice, the *MKL1*/*SRF* signaling pathway is also inhibited, and endogenous *MKL-1* nuclear translocation in cardiomyocytes is dysfunctional, with reduced expression of *SRF*, α-actinin and membrane association proteins, leading to the development of dilated heart disease[21]. Simple blockade of *SRF* expression in mouse myocardial tissue can lead to the development of DCM[22].

Zebrafish model is one of the ideal models to study the function of mutated genes in DCM, and harbored an enlarged heart, reduced shortening fraction during cardiac contraction and no cardiomyocyte proliferation, consistent with the characteristics of human DCM[23].

In this study, we performed whole-exome sequencing (WES) in consanguineous family DCM to unravel DCM-causing gene. Subsequently, functional studies of candidate DCM gene, *BICD2*, were conducted in zebrafish models to demonstrate the relationship between *BICD2* and DCM.

## Materials and Methods

### Patients and clinical evaluation

The proband and zir families from consanguineous family was recruited in 2011. In total 210 sporadic DCM patients who hospitalized at the Cardiology Department of Sichuan People’s Hospital from 2014 to 2017, were enrolled for further *BICD2* variant validation. This study was approved by the Institutional Research Committee of Sichuan Provincial People’s Hospital. Written informed consent was obtained from all subjects participating.

### Genomic DNA preparation

Peripheral blood from DCM cases was collected into EDTA anticoagulant tubes, and genomic DNA was extracted using a blood DNA extraction kit according to the manufacturer’s protocol (Qiagen, Germantown, MD, USA).

### Exome sequencing and bioinformatics analysis

To identify additional genes for DCM, we applied WES to search for potential genetic variants in DCM family without pathologic variants in known DCM genes. WES was performed on four members of family 1(the proband, sibling of the proband, parents of the proband). The normal human population database consisted of The Thousand Genomes Project (http://browser.1000genomes.org), ESP6500SI-V2 (http://evs.gs.was-hington.edu/EVS), ExAC Human Exome Integration Database (http://exac.broadinstitute.org/) and the sequencing company’s internal database. All variants were annotated with ANNOVAR software (version 2014).

### Quantitative RT-PCR

Total RNA was isolated from cells using the Qiagen RNeasy Mini kit. In parallel, we analyzed the mRNA concentration of the housekeeping β-actin as an internal control for normalization. The real-time monitoring of the PCR reaction, the precise quantification of the products in the exponential phase of the amplification and the melting curve analysis were performed with the Bio-Rad CFX Manager software, following recommended instructions of the manufacturer.

### Immunocytochemistry

Heart tissue fixed with 4% paraformaldehyde was dehydrated by an automatic dehydrator, embedded, and sectioned as the following protocols. Firstly, the dewaxed sections were placed in a staining vat with 3% methanolic hydrogen peroxide at room temperature for 10 min. Secondly, PBS wash 3 times (5 min each time). Then, immersing the sections in 0.01 M citrate buffer (PH 6.0), heating to boiling using Microwave Oven and then disconnecting, after an interval of 5 min, repeated once, after cooling, washed 2 times with PBS (5 min each time). Adding normal goat serum blocking solution dropwise, room temperature for 20 min. Adding primary antibody dropwise, overnight at 4°C. Adding biotinylated secondary antibody dropwise, 37°C for 30 min. PBS wash 3 times (5 min each time). Mixing the reagents of DAB Color Development Kit (K135925C, Beijing Zhongshan Jinqiao Biotechnology Co., LTD), and add dropwise to the sections at room temperature, for about 2 min, then wash with distilled water. Hematoxylin lightly re-stained, dehydrated, transparent, and sealed with neutral gum. Epifluorescence Images were acquired using an BA200 Trinocular Microscope (McAudi Industrial Group Co., Ltd).

### CRISPR/Cas9 *bicd2* knock out in zebrafish

Considering the conserved functional domain of the protein encoding by bicd2, we designed CRISPR sequence to target the genomic sequence corresponding after coding 80 amino acids, so as to disrupt the conserved structural domain of the BicD superfamily. And it is recommended that the CRISPR sequence be designed in Exon 2. Four guide RNA targeting the *bicd2* locus was designed following criteria of high on-target score and low off-target score. Zebrafish fertilized eggs were collected. Cas9 protein(a final concentration of 200 ng/ul) were mixed with sgRNAs (a final concentration of ∼ 80 ng/ul) and subsequently microinjected into zebrafish fertilized eggs (1 nl per embryo). Five embryos were taken when the injected embryos developed to 24 hpf, and genomic DNA templates were prepared under the following reaction conditions: 65°C for 30 min, 95°C for 5 min, 16°C for 1 min, 4°C for 2 min. PCR amplification products were cloned into pGEM-T Easy plasmid (5 μl total volume of reaction). The above 5 μl ligation product was transformed into 50 μl E. coli DH5α receptor cells (pfu≥108). Then coated in LB plates containing ampicillin (50 μg/ml) and incubated upside down overnight at 37°C. Positive clones were validated by Sanger sequencing. Experimental results confirmed that sgRNA1 and sgRNA2 could effectively guide Cas9 to target cleavage of *bicd2*.

*bicd2*-sgRNA1:

<u>GGUUGAGUGAACCUGGCCAU</u>GUUUUAGAGCUAGAAAUAGCAAGUUAAA AUAAGGCUAGUCCGUUAUCAACUUGAAAAAGUGGCACCGAGUCGGUGC UUUUUUU

*bicd2*-sgRNA2:

<u>GGAGUCGCUCAUCCUGGAGU</u>GUUUUAGAGCUAGAAAUAGCAAGUUAAA AUAAGGCUAGUCCGUUAUCAACUUGAAAAAGUGGCACCGAGUCGGUGC UUUUUUU

*bicd2*-sgRNA3:

<u>GUACUAUGAGCAGAGGGUGC</u>GUUUUAGAGCUAGAAAUAGCAAGUUAAA AUAAGGCUAGUCCGUUAUCAACUUGAAAAAGUGGCACCGAGUCGGUGC UUUUUUU

*bicd2*-sgRNA4:

<u>GCUCGAGGGCAAGGGUGGCC</u>GUUUUAGAGCUAGAAAUAGCAAGUUAAA AUAAGGCUAGUCCGUUAUCAACUUGAAAAAGUGGCACCGAGUCGGUGC UUUUUUU

### Zebrafish echocardiogram

The Vevo2100® Imaging System and Vevo Imaging Station (VisualSonics) was used to perform transthoracic echocardiography on 7-month-old zebrafish to examine indicators of ventricular function and ventricular size during systole and diastole with a 22–55 MHz (MS700) transducer probe. Before echocardiography, zebrafish were anesthetized with 0.02% tricaine (MS-222) to induce sedation without cessation of breathing. Anesthetized fish were then transferred into a glass beaker containing a weighted sponge with a groove. The fish was fixed on a sponge in the supine position. Echocardiography were collected and analyzed from the apical 2-chamber view. We typically completed image acquisition within 5 min of inducing anesthesia. After echocardiography, fish were placed in an aerated recovery chamber containing fresh system water without anesthetic and they recovered within 30 s to 2 min, and there were no deaths. The two-dimensional B-mode was used for measurement of heart rate, ejection fraction, fractional shortening, end-diastolic and end-systolic area, end-diastolic and end-systolic volume, cardiac output, and stoke volume. The left ventricular volumes are calculated by tracing the endocardial border manually at end diastole and at end systole (planimetry). B-mode imaging quality was further optimized by adjusting focal depth, gain, image width and depth.

### RNA-seq and data analysis

Total RNA from zebrafish hearts was isolated from cells using the Qiagen RNeasy Mini kit. Qubit2.0 Fluorometer was used to measure the quality and concentration of mRNA. Commercial sequencing companies sequenced our mRNA samples by Illumina Hiseq X and returned raw sequencing reads to us. Before further data analysis, we firstly checked raw data quality and removed reads with poor quality to get clean reads with high quality. Then all clean reads were mapped to zebrafish genome Assembly GRCz11 (https://hgdownload.soe.ucsc.edu/goldenPath/danRer11/bigZips/genes/danRer11.ensGene.gtf.gz) using HISAT (version 2.2.1) [24]. StringTie (version 2.2.0)[24] was used to quantify gene expression in different samples and FPKM values were extracted as the expression metric. EBSeq (version 1.34.0) [25] was utilized on raw reads matrix to find out differentially expressed genes. Genes with absolute fold change not less than 2, and adjusted P-value less than 0.05 were defined as differentially expressed genes.

### Statistical analysis

Statistical significance was determined using Student’s t test or Wilcox test and was assumed at P□<□0.05. Quantitative data are presented as the means□±□se, as indicated in the figure legends. Statistical analyses were performed using R 4.10.

## Results

### Identification of candidate variants for the consanguineous family with DCM

A consanguineous family with three members affected DCM (family-1) was recruited. The proband from family-1 was diagnosed with DCM (NYHA class II). The results of conventional and dynamic electrocardiograms suggested occassionally first-degree atrioventricular block, paroxysmal sinus tachycardia and complete left bundle branch block (Fig. 1A). Increased circulating brain natriuretic peptide(BNP) and mildly elevated troponin were also observed. A sibling of the proband was diagnosed with severe DCM, when ze presented with a short history of symptoms of congestive heart failure. Ze experienced a rapid deterioration and eventually died one year after surgery treatment. A sibling of the proband, IV-3, was diagnosed with DCM (NYHA class II) after presentation to clinic with respiratory distress. Ze had a left ventricle ejection fraction (LV EF) value of 55%, slightly dilated LV(a diameter of 52 mm in left ventricular). All three DCM patients were children of consanguineous parents, but none of the parents or other ancestors showed cardiac dysfunction.

**Fig. 1.**
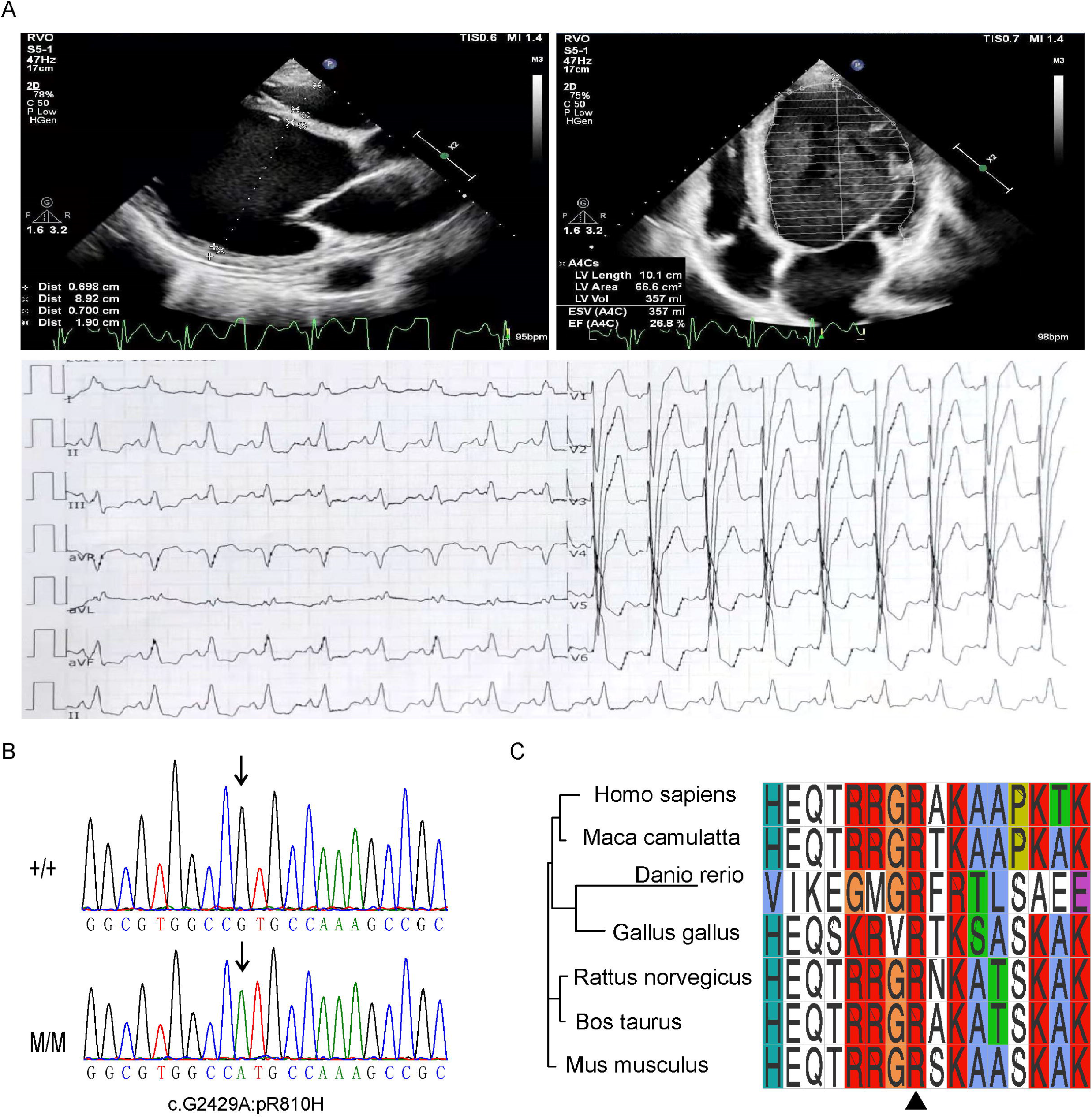
Family pedigree and clinical phenotypes. A. Echocardiography and electrocardiogram images of the proband showing dilated heart and decreased LVEF. Upper left panel: parasternal long-axis echocardiogram view of the left ventricle showing two-dimensional measurements of right and left ventricular wall thickness, septal thickness and right and left ventricular internal diameter. Upper right panel: apical four chamber echocardiogram view showing length, area, volume, end-systolic volume(ESV) and ejection fraction of the left ventricle. Lower panel: 12 lead electrocardiogram(ECG) showing paroxysmal sinus tachycardia and complete left bundle branch block. B. Sanger sequencing results indicating the genotypes of the family members. Arrows indicate the variant locus. C. Protein sequence alignment of amino sequences surrounding the *BICD2* variant with orthologues from H. sapiens to Danio Rerio. Note that the amino acid sequences surrounding the affected amino acid residues are highly conserved.

Whole-exome sequencing were performed on the consanguineous family members with DCM, but found no known DCM-causing gene mutations, we then turned to seek new candidate pathogenic genes. The analysis of high-throughput second-generation sequencing data proceeded as illustrated (Supplementary Fig. S1). In brief, we firstly filtered out nonharmful variations and acquired harmful variations for further analysis. Subsequently, common variants in East Asians, which defined as variations with population frequency ≥5% in the normal human population database consisting of The Thousand Genomes Project, ESP6500SI-V2, ExAC Human Exome Integration Database and in-house database, were further removed, leaving 14,208 variants for further selection. Considering autosomal recessive inheritance pattern of the consanguineous family, both DCM patients including the proband should be a homozygous affected individuals, and parents of the proband could be heterozygous carriers. And only fifty-six variants accorded with the autosome recessive genetic pattern were regarded as candidate disease-causing variants. Finally, fifty-one more variants were left out based on the American College of Medical Genetics and Genomics (ACMG) 2015 guidelines, resulting in five variants located at five separate genes, *BICD2, SERINC1, BVES, ADCY1* and *PAPPA* for further evaluations (Table S1).

*BICD2* is a dynein activating adaptor protein involved in microtubule-based minus-end-directed transport via docking the dynein motor proteins to appropriate cargos. Lower *LMNA* protein expression and *MKL-1*/*SRF* activity were observed in *Bicd2*-deficiency mice reported previously by Dick et al[20]. It is known that *LMNA* is the most common causative gene for DCM, accounting for 4%-8% of patients with DCM[26-28]. In addition, some individuals with spinal muscular atrophy caused by the *BICD2* ‘hot spot’ mutation, c.302C>T:p.Ser107Leu, surprisingly presented the heart failure associated symptom, exertional and supine dyspnoea[18]. Considering the possible interaction relationship between *LMNA* and *BICD2*, and the heart failure symptom that *BICD2* mutant patient exhibited, we hypothesized that the homozygous variant (exon7:c.G2429:p.R810H) in C-terminal region of *BICD2* (Table S1) would be a candidate variant that caused DCM in this family.

The missense variant of *BICD2* (exon7: c.2429G>A:p.Arg810His) (Table S1), segregated with the disease phenotype. Prediction score of the SIFT(sorting intolerant from tolerant) algorithm[29] was 0.003(Supplementary Fig. S2), and the variant was considered to be damaging to protein function. Sanger sequencing was further utilized to validate the variant (exon7: c.2429G>A:p.Arg810His) in DCM patients from family 1. Sanger results confirmed that both parents were heterozygous carriers and the two surviving DCM patients (the proband and sibling) were homozygous at this locus (Fig. 1B). Furthermore, the amino acid affected by this variant are highly evolutionarily conserved in not only H. sapiens but also Macaca mulatta, Bos taurus, rattus norvegicus, Mus musculus, Gallus gallus, and Danio rerio (Fig. 1C). It is well known that evolutionarily conserved sites in a multiple sequence alignment usually correspond to functionally important sites. Therefore, the missense variant in the evolutionarily conserved site (encoding amino acid R) could be harmful to protein function.

### Verification of three *BICD2* variants from 210 sporadic DCM cases

Patients hospitalized at the Cardiology Department of Sichuan People’s Hospital at least once for heart failure from 2014 to 2017 were enrolled in our functional research about *BICD2*. We further selected patients diagnosed with DCM and patients diagnosed with abnormal loading conditions, coronary artery disease, hypertension, valvular disease or congenital heart disease. Following the above criteria, we obtained in total 210 sporadic DCM patients for further *BICD2* mutant validation.

In 210 sporadic DCM cases, we performed full exon sequencing of *BICD2* and found one missense variant and two synonymous variants (Fig.2). The missense variant located in exon 2 (c.421G>A, p.Arg141Ser) (Fig. 2A) contributed to amino acid shifting from arginine to serine. Arginine is a basic amino acid and serine is an uncharged amino acid. Besides, algorithm prediction of functional impact suggested that it was a deleterious variant (Table S2). But no information about this variant could be acquire in normal population databases including The Thousand Genomes Project, ESP6500SI-V2, ExAC Human Exome Integration Database, gnomeAD, and ESP Exome sequencing project. The other two variants of *BICD2* were Synonymous (Table S2). These results support that *BICD2* may paly a significant role in DCM pathogenesis.

**Fig. 2.**
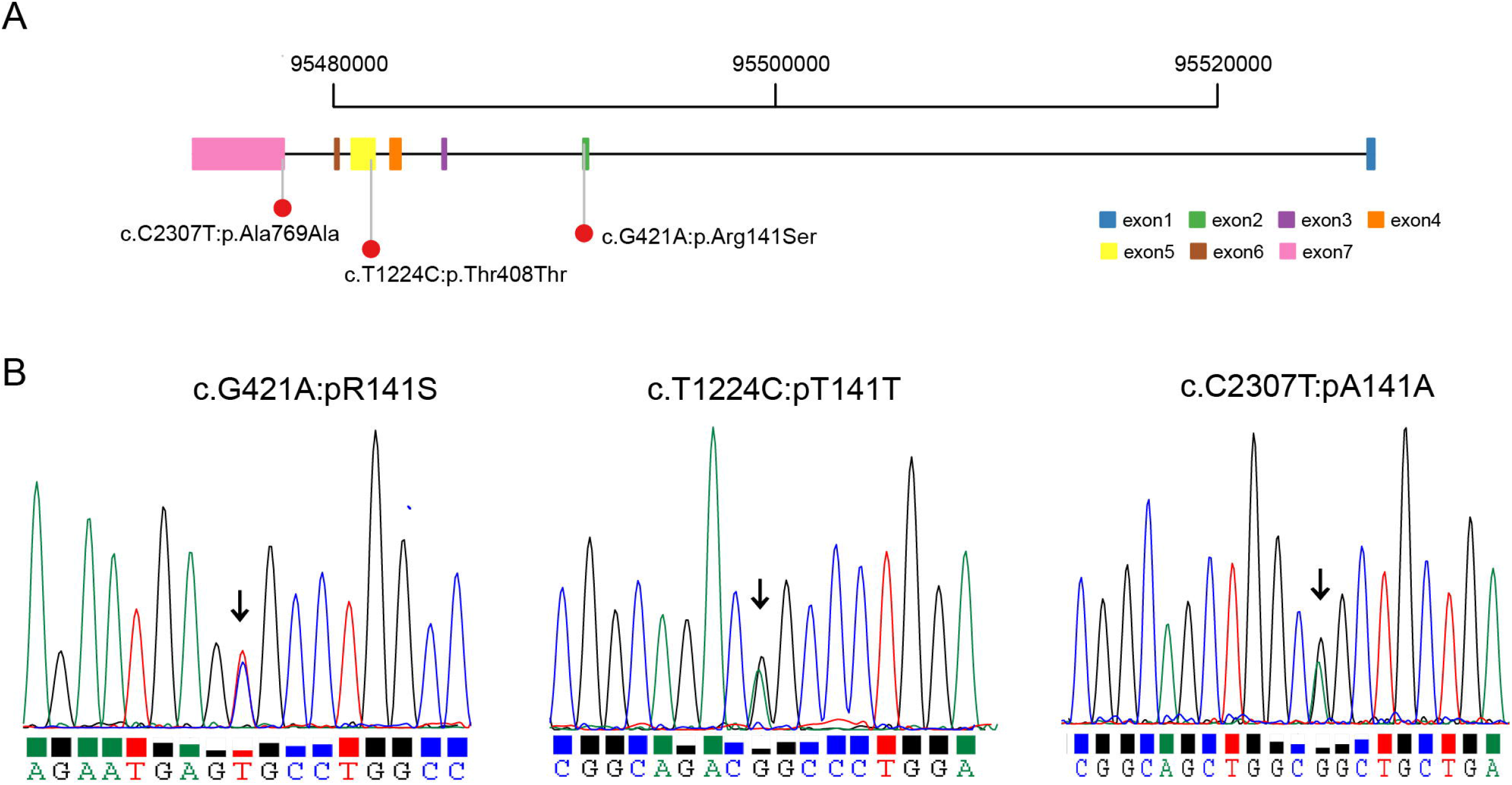
Validation of *BICD2* in 210 sporadic DCM patients. A. Three *BICD2* variants identified in 210 sporadic DCM patients. B. Sanger sequencing results of the missense variant.

### Expression status of *BICD2* in heart tissue

We explored to quantify expression of *BICD2* in heart of human as well as mouse and zebrafish. Real-time PCR results showed that *Bicd2* mRNA expressed in hearts of 1-week, 4-week old C57 mice, with increasing expression level upon age (Fig. 3A). Similar expression pattern was also observed in hearts of zebrafish and expression of *bicd2* measured by qPCR was much higher at stage of 7 months than that at stage of 5 months (Fig. 3B).

**Fig. 3.**
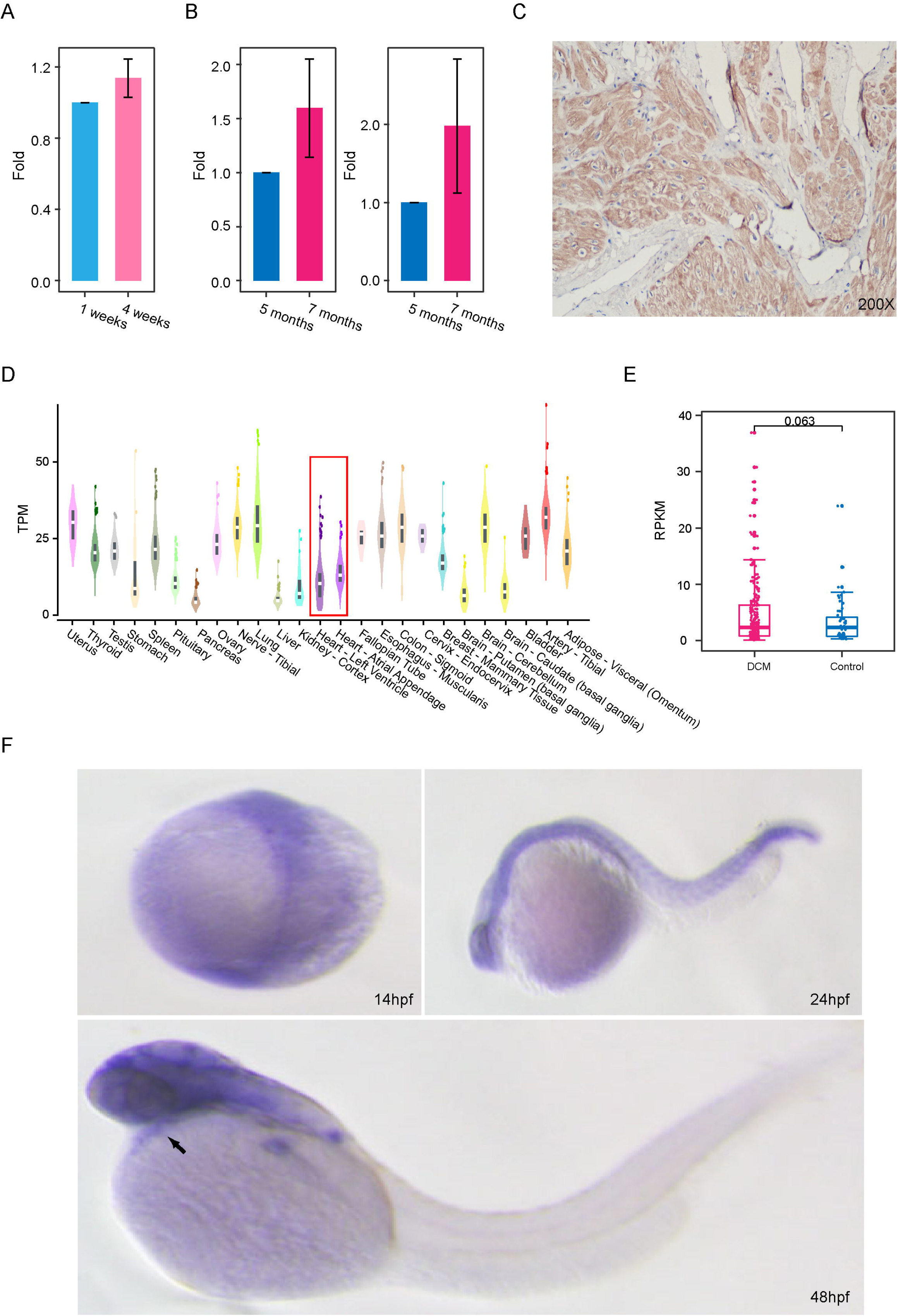
Expression of *BICD2* in hearts of human, mouse and zebrafish. A. qPCR results of *Bicd2* expression in hearts of C57 mouse suggesting an increasing trend over developmental stages(n=3). Data are represented as the mean ± SD. B. qPCR results of *BICD2* expression in hearts of zebrafish suggesting an increasing trend over developmental stages(n=3). Data are represented as the mean ± SD. C. Immunohistochemistry with anti-*BICD2* antibody of normal human heart showing ubiquitous staining. D. Bulk RNA-seq revealed mRNA expression (transcript per million fragments) of *BICD2* in multiple tissues from human. E. *BICD2* expression difference in scRNA-seq of cardiomyocytes from DCM(n=213) and control(n=52) deposited in GSE95140. Student t test, pvalue=0.063. F. In situ hybridization of *bicd2* in zebrafish revealed that *bicd2* expressed in hearts at embryonic stage.

Subsequently, we further detected the expression of *BICD2* at the protein level. Immunohistochemical staining of *BICD2* in normal heart tissue (left ear) supported its high expression in human(Fig. 3C). Moreover, *BICD2* bulk RNA-seq data from GTEx database confirmed its relatively higher expression in human heart (Fig. 3D), and scRNA-seq data of *BICD2* suggested that it mainly expressed in endothelial cells, fibroblasts, immune cells (Supplementary Fig. S3). Furthermore, published scRNA-seq (GSE95140) of cardiomyocytes from human hearts revealed that BICD2 expressed slightly higher in DCM and that in control (median RPKM of BICD2 in DCM and control is 2.33 and 2.28)(Fig. 3E), implicating a vital role of BICD2 in regulating DCM cardiomyocytes. Thus, we demonstrated that *BICD2* expressed in hearts of human, and mouse and zebrafish at both mRNA and protein levels.

We also conducted in situ hybridization of *bicd2* in zebrafish at all three embryonic stages to explore its expression status (Fig. 3F). At 14hpf, *bicd2* expressed in the axial midline cells of embryos. In detail, weak expression were observed in the somatic segment and strong expression were witnessed in the caudal notochord. At 24hpf, expression of *bicd2* was observed in eye, ventral part of the 4th ventricle, and hindbrain of zebrafish. At 48hpf, *bicd2* expressed in the retina, forebrain, midbrain and hindbrain regions, with strong expression in the pectoral fin bud base (Fig. 3F). Lateral and abdominal views of embryos showed that *bicd2* was weakly expressed in the heart region. Taken together, *BICD2*/*Bicd2*/*bicd2* expressed at heart of multiple species, implying its involvement in heart development and function.

### Knockout of *bicd2* lead to partial embryonic lethality in homozygotes and altered cardiac function

To further investigate the molecular mechanism of *BICD2* in the pathogenesis of DCM, we designed a *bicd2* knockdown assay in zebrafish to unravel associated phenotypic changes, and the experimental workflow is shown in Fig. 4A. We injected Cas9/sgRNA into embryos of F0 generation zebrafish, and further screened for *bicd2*-deficiency zebrafish in F0 adult zebrafish (Fig. 4A)(CRISPR target sequence in Table S3). Three fish were obtained, and they were mated with each other to further screened *bicd2* heterozygous F1 generations(Fig. 4A)(PCR primers for F1 mutant identification in Table S4). Details for generation of *bicd2*-KO zebrafish is depicted in Supplementary Fig. S4. In detail, we identified one type of mutation for F1 zebrafish, with 42-bp deletion (42bp deletion and 2bp insertion) at target sites, causing gene frame shift and induce early termination of the encoded protein. We then allowed the F1 generation fish to self-cross to get F2 generation fishes with three segregating genotypes (Fig. 4A). We observed the F2 generation fish in 2 different periods, i.e. embryonic stage and adult stage. In the embryonic period, we mainly observed the survival rate of embryos of different genotypes, and we measured the survival rate of embryos in three periods: 50hdf, 76hdf and 120hdf (Fig. 4A). Considering the insufficient number of homozygotes, we observed three batches of zebrafish at three different embryonic stages. At the adult stage, we performed echocardiographic measurements and transcriptome sequencing to compare *bicd2-*deficient to wild-type fish to delineate possible regulatory mechanisms underlying the cardiac dysfunction caused by *bicd2* (Fig. 4A).

**Fig. 4.**
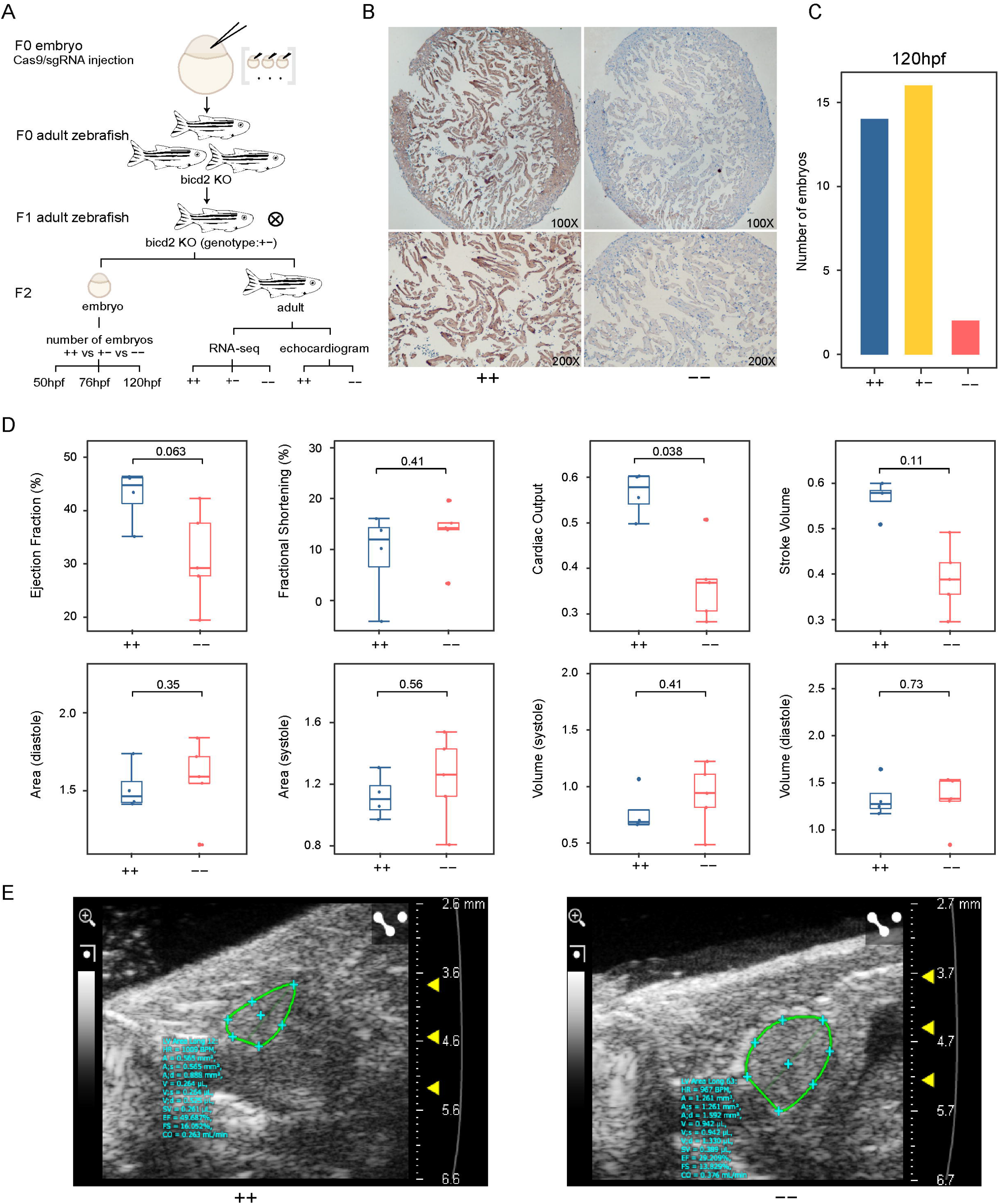
DCM associated phenotypes in *bicd2*-deficient zebrafish. A. Flow chart of *bicd2* zebrafish models for functional experiment. B. Immunohistochemistry with anti-*BICD2* antibody of zebrafish hearts showing the absence of staining in *bicd2*--fishes and ubiquitous staining in wild-type fishes. C. Number of embryos in homozygotes(n=2), heterozygotes(n=16) and wild-type fishes(n=14) at embryonic stage of 120hdf. D. Area, volume, Ejection Fraction, Fractional shortening, cardiac output, and stroke volume difference in homozygotes(n=5), and wild-type fishes (n=4). E. Echocardiogram plot showing ejection fraction difference in a homozygote and a wild-type fish.

We firstly examined *bicd2* by immunostaining to check whether *bicd2* efficiently knocked out from zebrafish hearts. And immunostaining images of the *bicd2*-deficiency fish showed lower *bicd2* expression than the wild-type group (Fig. 4B). We then analyzed number of viable embryos and found that the number of viable embryos was much lower in homozygotes than that in the other two groups, at all three time points (Fig. 4C, Supplementary Fig. S5). At 50hpf, there was only one homozygous embryo survived (Supplementary Fig. S5) (Table S5), while the number for heterozygotes and wild-type fish were twenty-two and thirteen(Supplementary Fig. S5) (Table S5), respectively. The proportion of viable homozygous embryos was 2.78%, far less than the theoretical ratio 25%. Similar trend was observed at both 76hpf and 120hpf, and the proportion of viable homozygous embryos was 15% and 6.25%(Fig. 4C, Supplementary Fig. S5) (Table S5), respectively. The embryonic viability data we observed for different genotypes suggests that *bicd2* plays a vital role in zebrafish embryogenesis and supports the conclusion that *bicd2* partially lead to death. We also examined heart rate of these three genotype groups at 50hdf, 76hdf and 120hdf, but we found no significant difference (Supplementary Fig. S6) (Table S6). Besides, no obvious phenotype was observed during embryo development period (images not shown). Then we turned to analyze the corresponding metrics of cardiac function at adult stage.

Electrocardiography was performed on wild-type and homozygous adult fish at seven months. In zebrafish echocardiographic studies, ventricular chamber size has mainly been assessed using volume and area. We therefore measured two-dimensional end-diastolic area (VAd) and end-systolic area (VAs), and three-dimensional end-diastolic volume (EDV), and end-systolic volume (ESV) in both wild-type and homozygous zebrafish. Compared with wild-type zebrafish, the *bicd2*--fish, showed slightly larger value in both VAd and VAs (Fig. 4D, Table S7). EF was much lower in the *bicd2*-deficiency group than that in wild-type group (Wilcox test, P-value = 0.063) (Fig. 4D, Table S7). It is well known that EF is a measurement of a percentage of how much blood the left ventricle pumps out with each contraction, and calculated as the ratio of amount of blood pumped out to amount of blood in chamber. EF< 45% is a diagnosis measurement for DCM[3]. And in our studies, most homozygotes presented EF<45%, supporting that *bicd2* was crucial for cardiac function. What’s more, cardiac output was significantly lower in the *bicd2*--fish than that in wild-type fish (Wilcox test, P-value <0.05) (Fig 7A, Table S7). In addition, the *bicd2*--fish showed lower stroke volume (Fig. 4D, Table S7). Echocardiographic images represented zebrafish hearts with two different genotypes were extracted from echocardiogram and were comparably listed (Fig. 4E). In summary, these results indicate that *bicd2* knockout may cause abnormal contraction of the heart.

### RNA-seq of bicd2-deficient zebrafish revealed cardiomyocyte transcriptome shift

We conducted transcriptome sequencing in hearts of *bicd2* knockout zebrafish and wild-type zebrafish to examine the shifts in their transcriptional profiles. Consistent with expectations, *bicd2* expression was significantly lower in the knockout group than in the wild-type group (Table S8). Gene Set Enrichment Analysis (GSEA)[30] based on ranking of all expressed genes could discern more telltale biological clues than enrichment analysis solely focused on differential expressed genes (DEGs). Therefore, we firstly utilized GSEA to unravel significantly enriched gene sets. Compared to the wild-type group, the bicd2-homozygous group showed enrichment for cardiopathy-related signaling pathways and metabolic pathways, for example KEGG_CARDIAC_MUSCLE_CONTRACTION, KEGG_CALCIUM_SIGNALING_PATHWAY, KEGG_OXIDATIVE_PHOSPHORYLATION, KEGG_GLYCOLYSIS_ GLUCONEOGENESIS, KEGG_CITRATE_CYCLE_TCA_CYCLE and KEGG_PEN -TOSE_PHOSPHATE_PATHWAY(Fig 5A), as well as pathways associated with nervous system disease (Table S9). The results accord with previous functional exploration of *Bicd2* in mouse, which proved a causal role of *Bicd2* in neuronal disorders, a dominant mild early onset form of spinal muscular atrophy[20]. Besides, pathways relate to energy metabolism in the mitochondria like oxidative phosphorylation (OXPH) and tricarboxylic acid cycle (TCA), are common characteristics of distinct heart failure demonstrated previously [31, 32]. Interestingly, many pathways (6 out of 14, 42.86%) (Fig. 5B) increased in both bicd2 homozygous and heterozygous groups, including KEGG_CALCIUM_SIGNALING_PATHWAY and KEGG_GLYCOLYSIS_GLUCONEOGENESIS. Only 3 common pathways (Fig 5C) decreased in those two groups, implicating different transcriptome atlas between homozygous and heterozygous groups.

**Fig. 5.**
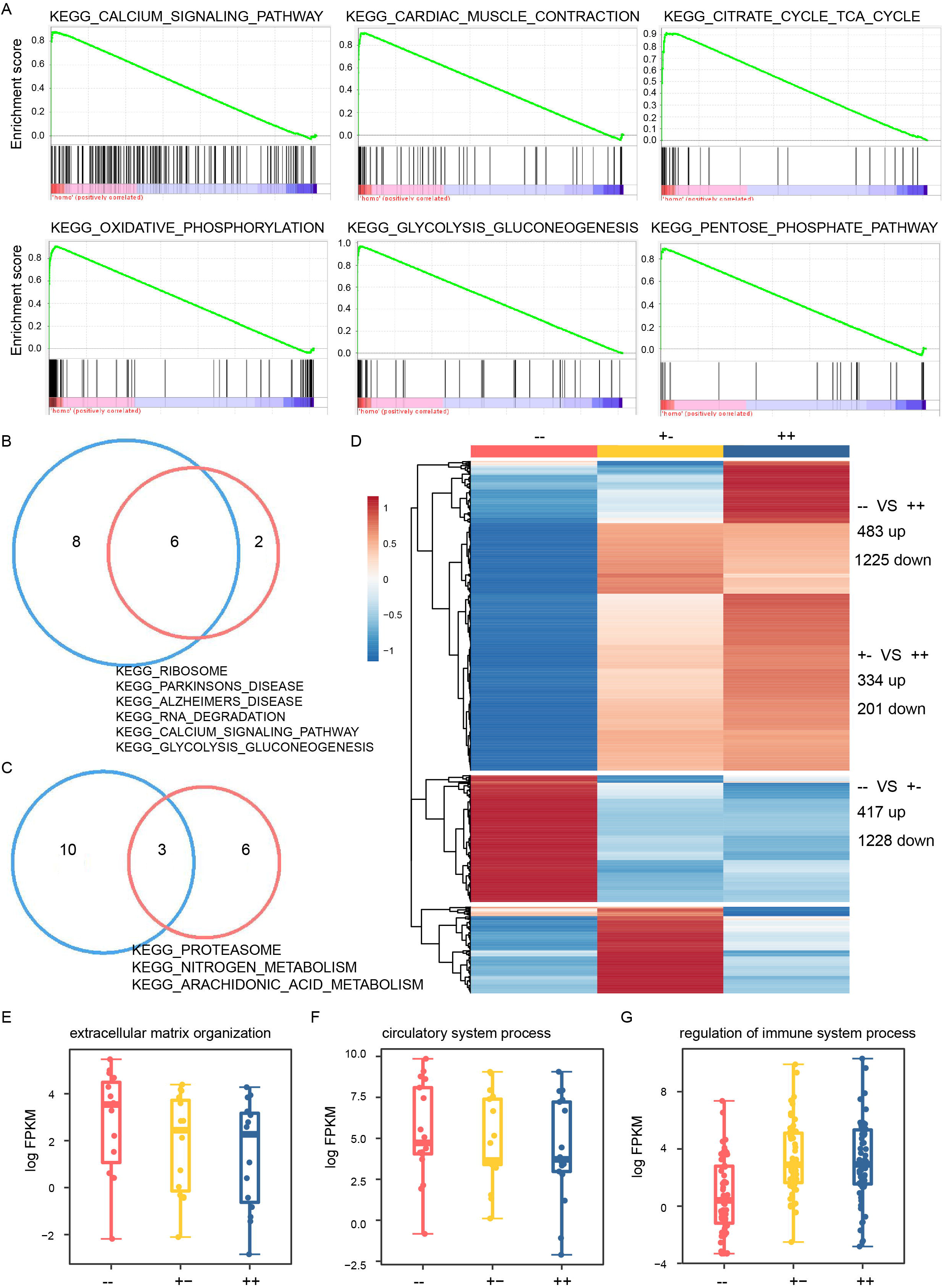
*bicd2*-deficient hearts exhibited cardiac transcriptome shift. A. GSEA plots of six KEGG pathways showing higher expression in *bicd2*--zebrafish. B. Venn diagrams showing enriched KEGG pathways increased in the *bicd2*-deficiency groups. C. Venn diagrams showing enriched KEGG pathways decreased in the *bicd2*-deficiency groups. D. Differential expressed genes in the two *bicd2*-deficiency groups. Cells are filled according to z-score: red indicates higher (activated), blue indicates lower (inhibited). E. Differential expressed genes in extracellular matrix organization. Cells are filled according to z-score: red indicates higher (activated), blue indicates lower (inhibited). F. Differential expressed genes in circulatory system process. Cells are filled according to z-score: red indicates higher (activated), blue indicates lower (inhibited). G. Differential expressed genes in regulation of immune system process. Cells are filled according to z-score: red indicates higher (activated), blue indicates lower (inhibited).

We then extract DEGs to further delineate biological meaningful pathways involved in *bicd2* gene regulatory network. Same as expected, the homozygotes displayed the largest change in all three groups, and in total 1708 DEGs (483 up-regulated genes, 1225 down-regulated genes, Fig. 5D, Table S10). Surprisingly, difference between the two *bicd2*-KO groups was much bigger than that between heterozygotes and the wild-type fish. In detail, there were 1645 DEGs (417 up-regulated genes, 1228 down-regulated genes, Fig 5D, Table S11) between homozygotes and heterozygotes, while 535 DEGs (334 up-regulated genes, 201 down-regulated genes, Fig 5D, Table S12) between heterozygous and wild-type groups. The top 15 enriched GO pathways based on 483 up-regulated and 1225 down-regulated genes in the homozygous group were displayed separately (Supplementary Fig. S7-8). Genes increased in the homozygous group mainly consisted of blood vessel development, extracellular matrix organization, cell-cell adhesion, carbohydrate metabolic process and circulatory system process (Supplementary Fig. S7). However, Genes decreased mainly related to the immune system, including regulation of immune system process, response to biotic stimulus, leukocyte activation (Supplementary Fig. S8).Notably, increased expression of genes encoding extracellular matrix proteins in DCM patients was previously demonstrated by microarray analysis[31]. In our study, extracellular matrix genes(Fig. 5E)(Supplementary Fig. S9) like *col4a2, col4a4, col5a1, col8a1b, col18a1b*, and genes of circulatory system process (Fig. 12) showed higher expression in the homozygous group than that in wild-type. Genes related to regulation of immune system process (Fig. 11) like *cd74b* and *cd79a* decreased in the two *bicd2*-KO groups, implying activation of the immune system after knocking out *bicd2* in zebrafish.

Fifty-one genes with human genetic evidence were proposed as causing susceptibility to inheritable DCM by previous studies[6]. Ortholog retrieval in the zfin data report (http://zfin.org/downloads/file/human_orthos.txt) and Ensembl (http://www.ensembl.org/index.html) identified zebrafish homologues for 48 of these genes(Table S13). Among these 48 genes, 28 had a single orthologue in zebrafish (Supplementary Fig. S 12, Table S13). 8 out of 28 orthologs including *bag3, nebl, psen2, jph2, dtna, psen1, gatad1* and *plekhm2* showed the lowest expression in the homozygous group(Supplementary Fig. S12). On the contrary, 10 out of 28 orthologs including *mybpc3, lama4, nppa, tnni3k, cmlc1, nexn, lrrc10, csrp3, abcc9, tcap* showed the highest expression in the homozygous group(Supplementary Fig. S12). There were also 20 human DCM genes that harbored multiple orthologues in zebrafish (Supplementary Fig. S13).

## Discussion

In this study, whole-exome sequencing of a consanguineous family with 3 DCM patients revealed a missense mutation (exon7: c.2429G>A: p.Arg810His) in *BICD2* was the corresponding pathological variant. Consanguineous couples of the family were both heterozygous carriers, and two DCM patients were homozygous genotype at this locus. Harmful effect of this mutation on amino acids were supported by algorithms, and the amino acid affected by these three variants are highly evolutionarily conserved in multiple species including Mus musculus, and Danio rerio, facilitating the functional exploration of the variant in mouse and zebrafish. Besides, genetic screening of *BICD2* in 210 sporadic DCM cases revealed three more variants of *BICD2*, further demonstrating the significance of *BICD2* involved in DCM.

Most previously reported familiar DCM cases are caused by pathogenic variants inherited in autosome dominant pattern[5]. Many DCM-linked genes encode proteins of the sarcomere, costamere, Z band, and nuclear membrane[5]. Currently more than 250 genes spanning >10 gene ontologies have been suggested contributing to inherited DCM[6]. However, heritability of DCM could not all be explained by variants discovered so far. In a consanguineous family we collected in clinical, the pathological variant for DCM was searched and a novel DCM-linked gene *BICD2* was discovered. Notably, the inheritance mode of DCM in the consanguineous family was autosome recressive pattern, suggesting loss of function caused the illness.

*BICD2* is a dynein activating adaptor protein that plays a critical role in microtubule-based minus-end-directed transport[13, 14]. *Bicd2*-KO mice showed significantly lower *LMNA* expression at protein level, decreased *MKL-1*/*SRF* activity, and significantly downregulated expression of its downstream α-actin, α-coactin and membrane association proteins, and microtubulin[20], thus affecting cardiovascular system development.

Furthermore, we conducted functional experiments in mouse and zebrafish to verify the relationship of *BICD2* and DCM. Real-time PCR results confirmed that *Bicd2* mRNA expressed in hearts of C57 mice and zebrafish. Immunostaining result showed that *BICD2* expressed in hearts of clinical patients at the protein level. Further *bicd2* knockdown assay in zebrafish revealed DCM-linked phenotypic changes. The survival rate of embryos in three periods, 50hdf, 76hdf and 120hdf revealed that *bicd2* is embryonic lethal, which is consistent with observations in *Bicd2*-KO in mice [20]. Different Ejection Fraction(EF) values between *bicd2*-deficiency and wild-type fishes was detected by echocardiography. RNA-seq of *bicd2*-KO revealed DEGs enriched in pathways relate to energy metabolism in the mitochondria like oxidative phosphorylation (OXPH) and tricarboxylic acid cycle (TCA), which are common characteristics of distinct heart failure demonstrated previously.

In this study, we identified a nonsynonymous pathological variant in *BICD2* from a consanguineous family, and we further found other *BICD2* variants associated with DCM, implying the significance role of *BICD2* played in DCM. We then demonstrated that deletion of *bicd2* in zebrafish decreased LVEF and altered energy metabolism in the mitochondria, implicating cardiac function dysfunction. Our study shows that *BICD2*, an adapter protein linking the dynein motor complex to various cargos, is a novel DCM candidate gene and conceptually expands our horizons regarding pathogenesis of DCM. To the best of our knowledge, this is one of the few reports of consanguineous family in the cardiovascular disease DCM, as evidenced by WES data, echocardiography, and zebrafish models.

However, we temporarily do not prove the direct relationship between the exact *BICD2* variant (exon7:c.G2429:p.R810H) and phenotypes associated to enlarged heart. We will continue to explore the cardiac dysfunction led by the *BICD2* variant. In addition, we will continue to validate the presumably pathologic variant in *BICD2* on more DCM patients in the future to provide additional evidence.

## Conclusions

Our study shows that *BICD2*, an adapter protein linking the dynein motor complex to various cargos, is a novel DCM candidate gene and conceptually expands our horizons regarding pathogenesis of DCM.

## Data Availability

All data produced in the present work are contained in the manuscript

## Abbreviations

DCM: dilated cardiomyopathy
WES: whole-exome sequencing
BICD2: Bicaudal-D2
LV: left ventricular
RNA-seq: RNA-sequencing
EF: ejection fraction
FS: fractional shortening
PAC: frequent premature atrial contractions
NYHA: the New York Heart Association
ACMG: the American College of Medical Genetics and Genomics
DEG: differentially expressed genes

## Authors’ contributions

Kai Luo: Conception and design, Collection and assembly of data, Data analysis and interpretation, Manuscript writing; Chenqing Zheng, Rong Luo: Provision of study material, Data analysis and interpretation; Xin Cao, Huajun Sun: Provision of study material, Assembly of data; Huihui Ma, Jichang Huang, Yang Xu: Responsible for manuscript writing and revision of the manuscript; Xiushan Wu: Collection and assembly of data, Data analysis and interpretation, Final approval of manuscript; Xiaoping Li: Conception and design, Manuscript writing, Financial support, Final approval of manuscript. Xiaoping Li is the guarantor of this work and, as such, had full access to all the data in the study and takes responsibility for the integrity of the data and the accuracy of the data analysis. All authors read and approved the final manuscript.

## Data Availability

The datasets used during the study are available are available within the article and its Supplementary materials.

## Acknowledgements

The authors would like to thank all patients who provided samples used in this study.

