## Supplemental Figure & Table for "Identification and Functional Characterization of Bicaudal-D2 As A Candidate Disease Gene in Autosomal Recessive Consanguineous Family with Dilated Cardiomyopathy": Supplementary Figure v3.pdf

### Supplementary Figures

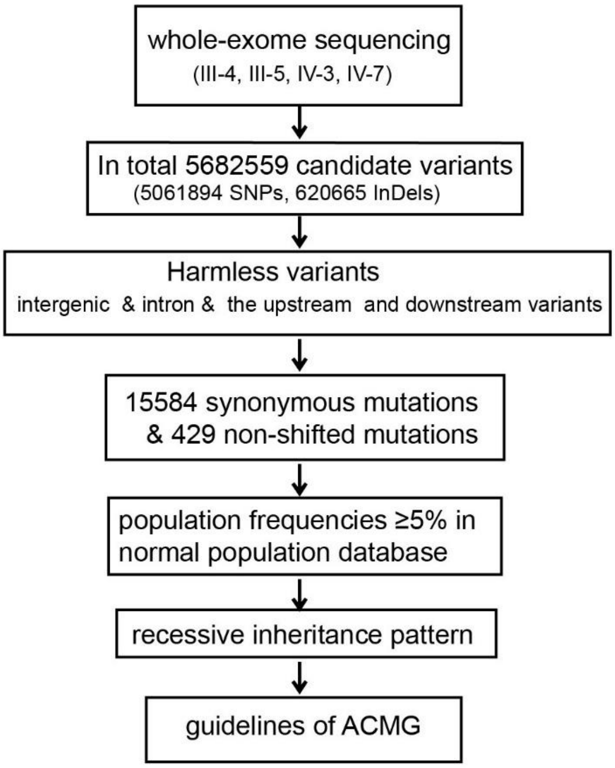


Supplementary Figure 1. Workflow of whole-exome sequencing analysis for the consanguineous family with dilated cardiomyopathy.

| PROTEIN SEQUENCE CHANGE | | | | | | PROVEAN PREDICTION | | | | SIFT PREDICTION | | | |
|---|---|---|---|---|---|---|---|---|---|---|---|---|---|
| INPUT | CODON_CHANGE | POS | RESIDUE_REF | RESIDUE_ALT | TYPE | SCORE | PREDICTION (cutoff=-2.5) | #SEQ | #CLUSTER | SCORE | PREDICTION (cutoff=0.05) | MEDIAN_INFO | #SEQ |
| 9,95477575,C,T | GGC C[G/A]T GCC | 810 | R | H | Single AA Change | -2.27 | Neutral | 169 | 30 | 0.003 | Damaging | 3.15 | 96 |

Supplementary Figure 2. Algorithm prediction for the *BICD2* missense variant (c.2429G>A:p.Arg810His) identified in DCM patients from the consanguineous family.

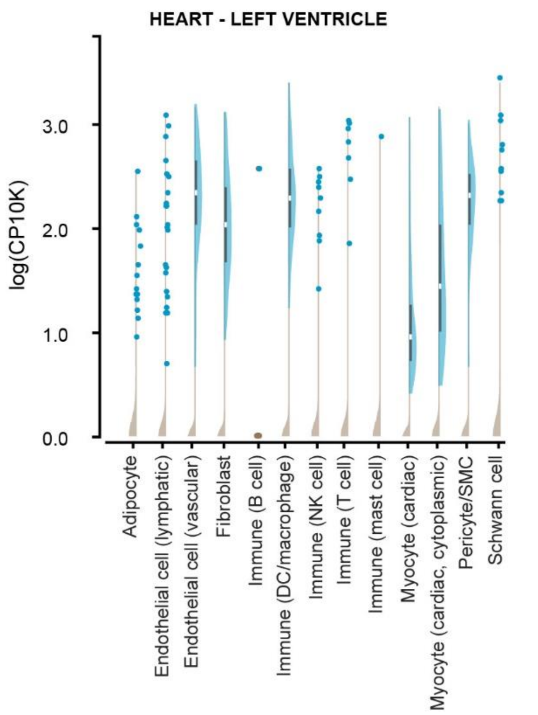


Supplementary Figure 3. scRNA-seq of the human heart unraveled *BICD2* mRNA expression in different cell types.

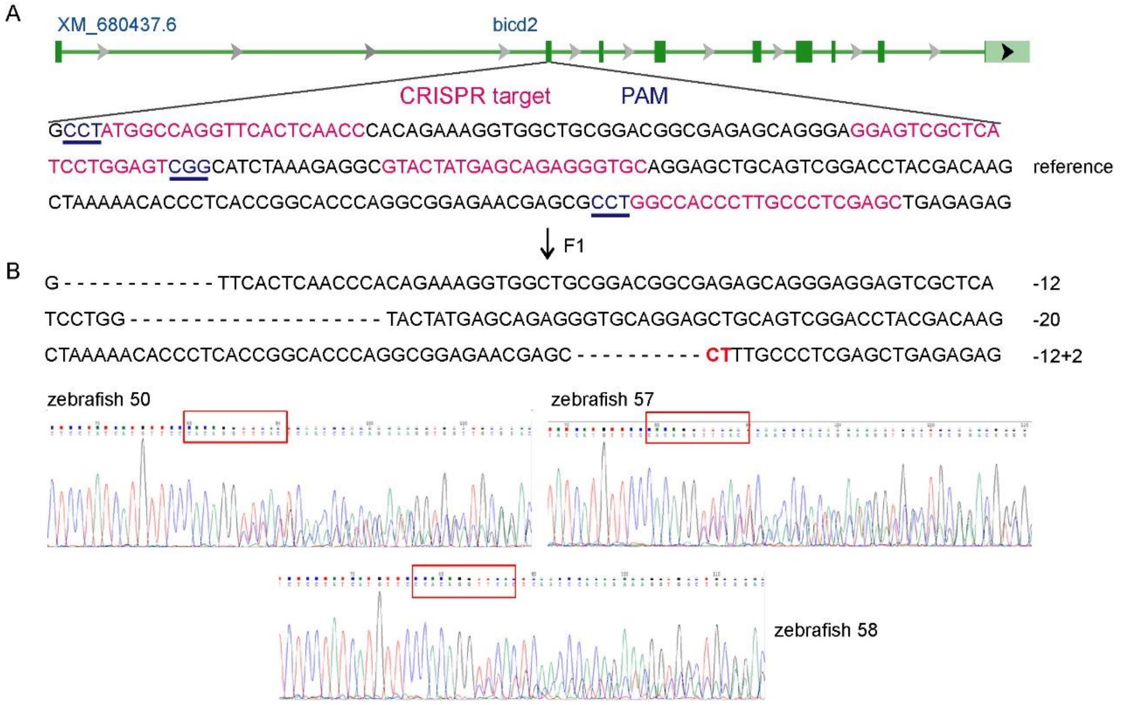


Supplementary Figure 4. Generation of bicd2 knockout zebrafish.

A. The target site of the CRISPR/Cas9 system, which was designed at the exon 2 of the bicd2 gene, consists of the red colored CRISPR target sequence and the CCT/CGG/CCT protospacer-adjacent sequence. We identified 3 heterozygous mutant adult zebrafish.

B. Sequencing validation of the F1 heterozygous mutant adult zebrafish (3 fish with the same mutant genotype).

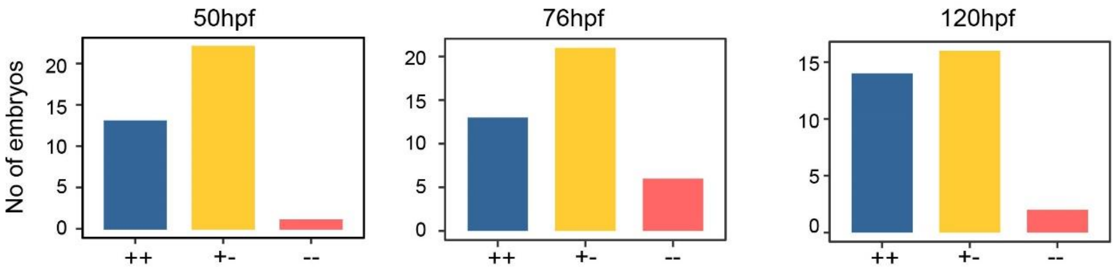


Supplementary Figure 5. Number of embryos in three different zebrafish groups with genotype of wild-type, heterozygous and homozygous

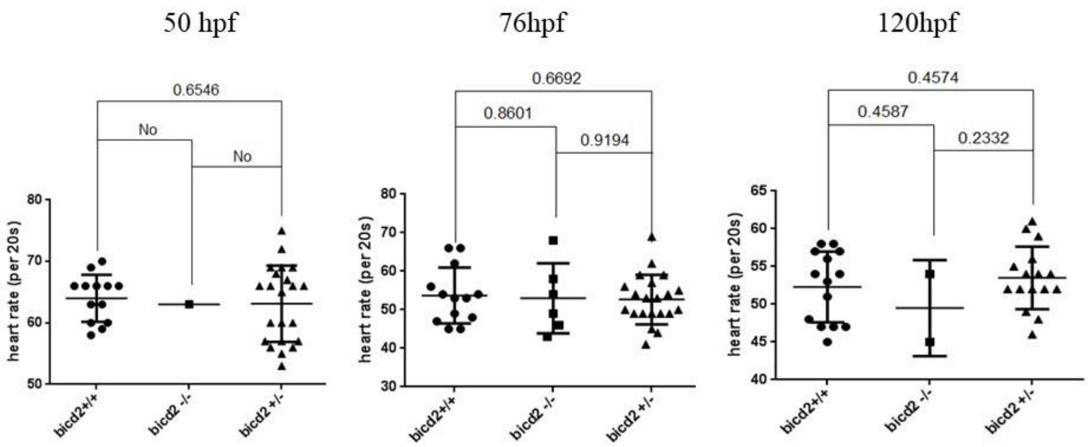


Supplementary Figure 6. Heart rate of embryo zebrafish in three different zebrafish groups with genotype of wild-type, heterozygous and homozygous

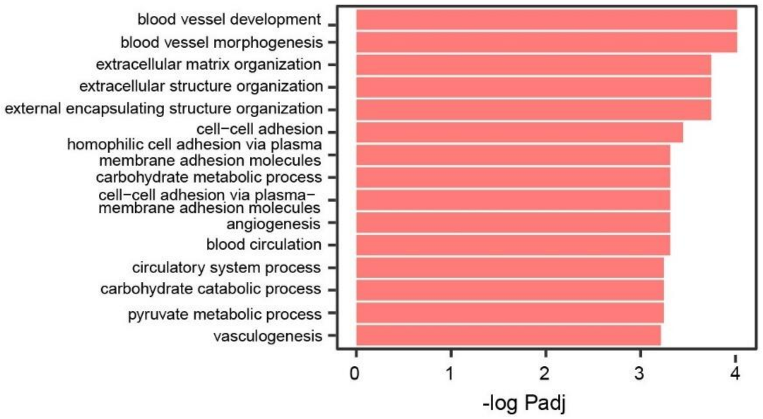


Supplementary Figure 7. The top 15 GO biological pathway increased in *bicd2* homozygous group.

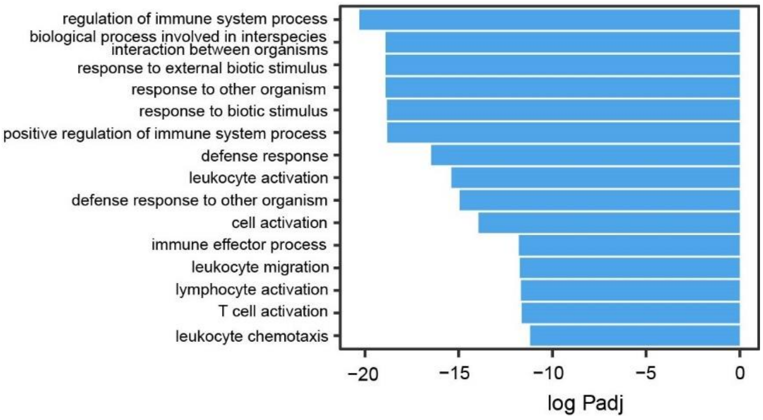


Supplementary Figure 8. The top 15 GO biological pathway decreased in *bicd2* homozygous group.

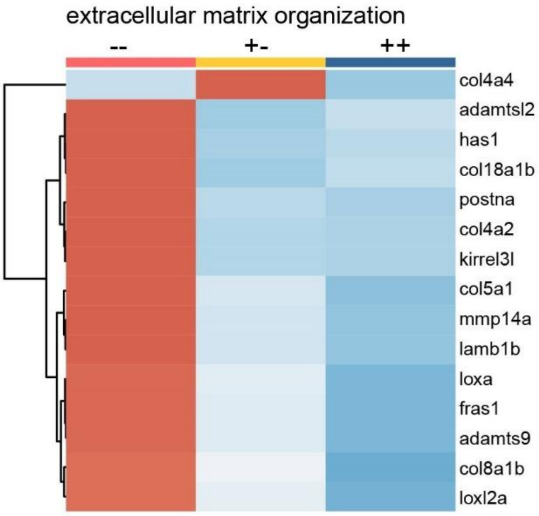


Supplementary Figure 9. Differential expressed genes in extracellular matrix organization.

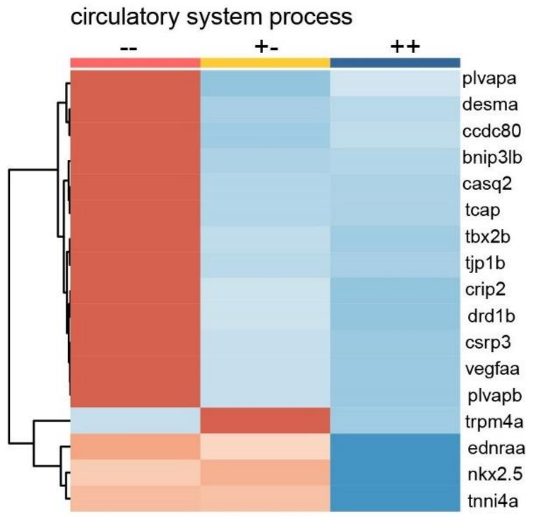


Supplementary Figure 10. Differential expressed genes in circulatory system process

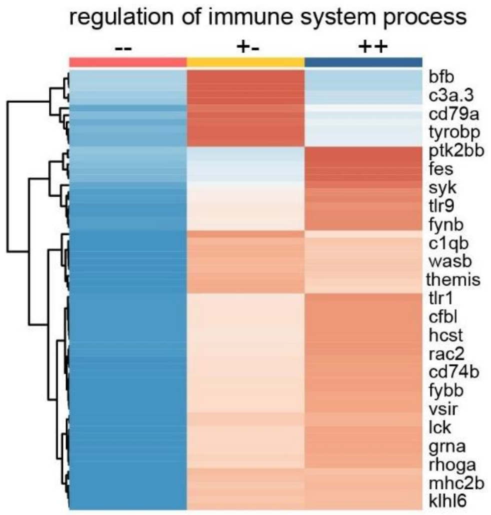


Supplementary Figure 11. Differential expressed genes in regulation of immune system process.

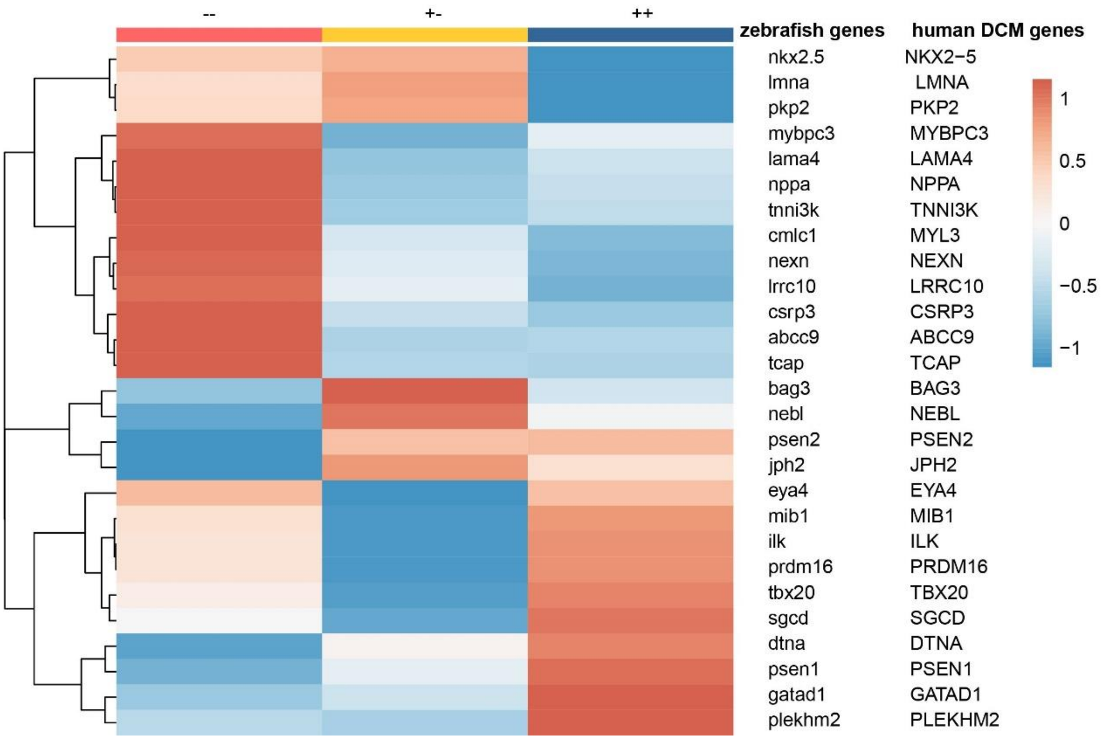


Supplementary Figure 12. Expression of zebrafish homologues of 20 human DCM associated genes with unique homologues in *bicd2*-deficiency zebrafish.

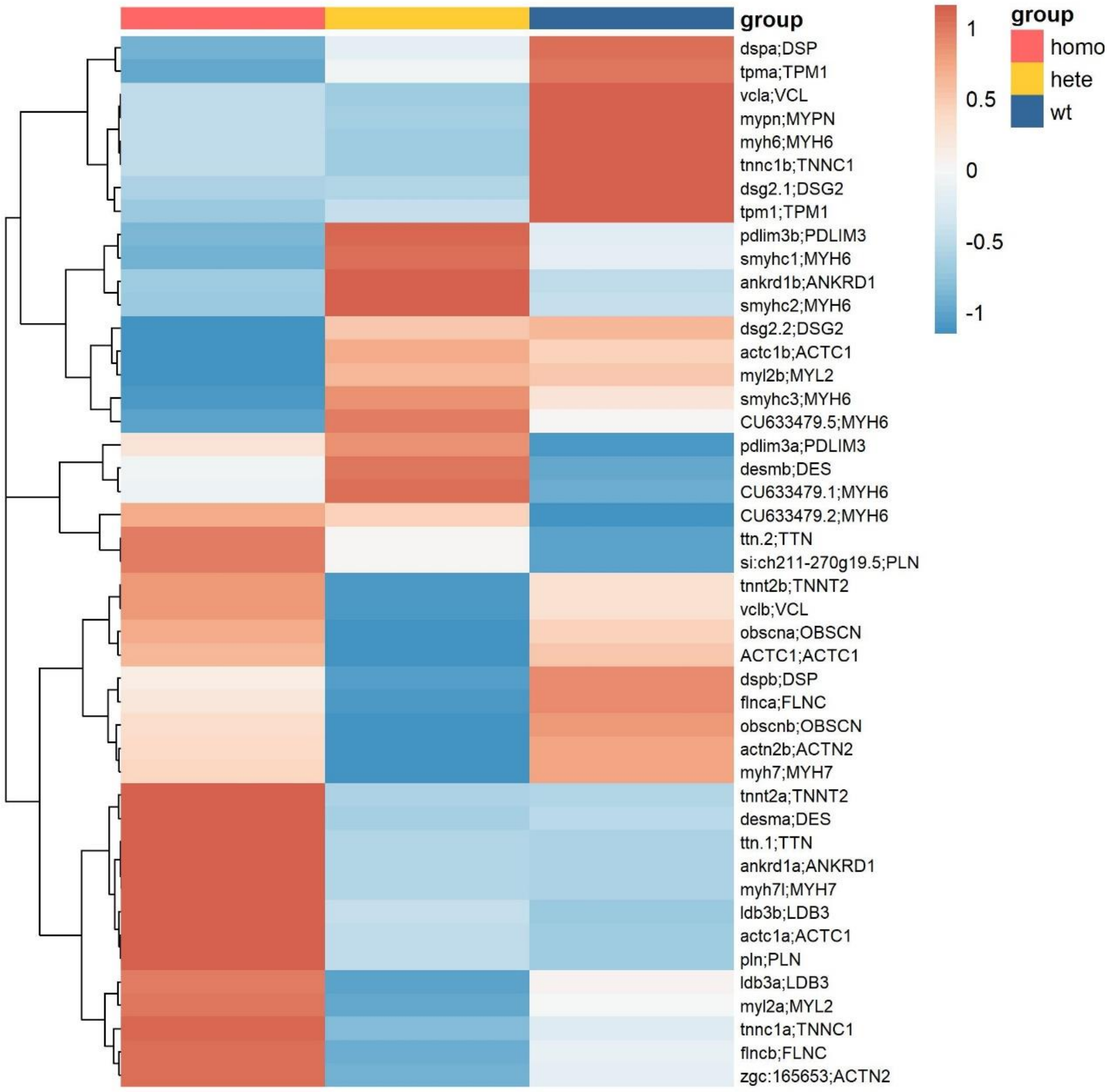


Supplementary Figure 13. Expression of zebrafish homologues of 20 human DCM associated genes with multiple homologues in bicd2-deficiency zebrafish.
